# Prediction of Major Clinical Endpoints in Atrial Fibrillation at Primary Care Level using Longitudinal Learning Stances

**DOI:** 10.64898/2026.03.26.26349389

**Authors:** Henrique Anjos, Ana Lebreiro, Cristina Gavina, Rui Henriques, Rafael S. Costa

## Abstract

Atrial fibrillation (AF) is the most prevalent cardiac arrhythmia worldwide and is strongly associated with increased risks of stroke, heart failure, and mortality. Traditional methods to predict AF and prognostic its associated risks often fail to capture the full complexity of AF patterns, limiting their predictive accuracy. In spite of the improvements achieved by machine learning (ML) techniques, state-of-the-art AF-focused predictors do not generally incorporate longitudinal data, reducing their capacity to model the dynamic and evolving nature of individual behaviors and physiological indicators over time. The absence of a longitudinal perspective restricts understanding of how AF risk develops and changes across prognostic windows. This study addresses these limitations by developing superior ML models tailored to predict adverse events within a longitudinal Portuguese cohort of individuals with AF. The work targets six clinical endpoints: stroke, all-cause death, cardiovascular death, heart failure hospitalizations, inpatient visits, and acute coronary syndrome. The predictors yielded an AUC of 0.65 for 1- year stroke prediction, outperforming CHA_2_DS_2_-VASc (0.59). For all-cause mortality prediction, the models achieved an AUC of 0.78 against the 0.72 reference of GARFIELD-AF. In addition to predictive advances, the study identifies determinants of AF-related risks and introduces a prototype decision-support tool for clinical use.

## 1 Introduction

Atrial Fibrillation (AF) is the most common cardiac arrhythmia worldwide, and its occurrence associated with a significant risk increase of stroke, heart failure, and mortality [10]. Studies estimate that around 9 million individuals in the European Union (EU) were affected by AF in 2010, and the number is projected to double by 2060 [24]. In Portugal, 4070 deaths were attributable to AF in 2010, corresponding to nearly 4% of all deaths [15]. Despite its prevalence and severe consequences, AF often goes undiagnosed until complications arise due to its episodic nature and the lack of consistent early-warning signs [8]. This diagnostic gap highlights the need for tools that can detect and predict AF and provide accurate prognostic assessment of its complications.

A plethora of traditional methods have been proposed for predicting AF and its complications that rely on point-based systems or traditional statistical models [20]. While these methods are functional, they often fail to capture the complexity of AF patterns [22]. In the last years, machine learning (ML) models have demonstrated superior performance compared to these traditional approaches [29]. However, the existing state-of-the-art ML methods generally suffer from two major drawbacks:

1. failing to incorporate longitudinal data, which limits their ability to analyze the progression of biometric and physiological indicators across varying prediction horizons.
2. underestimating the significance of integrating specific data modalities. Many approaches do not fully leverage the predictive value embedded in electrophysiological signals and neglect other critical factors such as risk behaviors, comorbidities, and drug regimen.

To address these challenges, this study leverages a longitudinal cohort of AF patients, using electronic health records from Unidade Local de Saúde de Matosinhos (ULSM) collected over the past decades to derive population-specific insights and to develop and optimize ML algorithms to accurately predict six AF-associated clinical end-points: stroke/systemic embolism (SE), all-cause mortality, cardiovascular death, heart failure hospitalizations, inpatient visits, and acute coronary syndrome. In parallel, a prototype clinical decision-support tool is designed to allow healthcare professionals to input patient data and visualize predictions intuitively. We also evaluate classical risk calculators, providing a comparative context, and establish a foundation for future development and clinical application.

This work is part of a broader initiative in collaboration with the Unidade Local de Saúde de Matosinhos (ULSM), which is expected to proceed in three phases: (1) prediction of complications in patients with AF; (2) prediction of AF onset in a case-control population; (3) incorporation of raw cardiac imaging and electrophysiological data to aid previous phases. This study focuses on phase 1, while also laying the groundwork for the following phases.

The main contributions of this work are the development of time-aware predictive models for AF-related outcomes, the systematic comparison of ML predictors against classical risk scores, the explainability-driven knowledge acquisition, and the design of a prototype clinical decision-support tool.

## 2 Related Work

Predictive scores for stroke risk in AF patients have been developed since the early 2000s [12]. The CHADS_2_ score is the primary stroke risk stratification scheme in patients with pre-existing AF and made part of the 2006 American College of Cardiology/American Heart Association/European Society of Cardiology (ACC/AHA/ESC) guidelines for nonvalvular AF [11]. This score assigns points based on the following risk factors: C - congestive heart failure (1 point), H - hypertension (1 point), A - age (1 point), D - diabetes mellitus (1 point), and S - prior stroke or transient ischemic attack (2 points). This score’s simplicity made it widely adopted; however, it did not account for certain risk factors, which led to the development of more refined scores.

The Birmingham 2009 schema, also known as CHA_2_DS_2_-VASc score [25] emerged as an improvement over CHADS_2_ and has been widely used since 2010 [30]. It provides a more comprehensive assessment of stroke risk in AF patients by incorporating additional risk factors, such as: V - prior vascular disease, A - age between 65 and 74 years, and Sc - sex category. This refinement significantly improved risk stratification, especially for patients at lower or intermediate risk [23].

In 2010, user-friendly HAS-BLED score was developed to assess 1-year risk of major bleeding in patients with AF, using data from the Euro Heart Survey on AF. The risk score gives points based on the following comorbidities: Hypertension, Abnormal renal/liver function, Stroke, Bleeding history or predisposition, Labile international normalized ratio, Elderly (*>*65 years), Drugs/alcohol concomitantly. The bleeding score achieved a consistent AUC of 0.72 in the derivation cohort across several subgroups.

In 2012, HAS-BLED bleeding risk score was experimented in predicting cardiovascular events and mortality in anticoagulated patients with AF, and was shown to be useful. However, a multivariate analysis was slightly better at predicting cardiovascular events and mortality, showing that HAS-BLED is not the most accurate score to predict these events [13].

In 2016, ABC (age, biomarkers, clinical history) stroke risk score was developed to predict 6-year stroke risk in AF patients with a Cox regression model. The ABC-Stroke score achieved higher AUCs than CHA_2_DS_2_-VASc in both the derivation cohort (0.68 vs. 0.62) and the external validation cohort (0.66 vs. 0.58). The score used biomarkers such as Troponin I and NT-proBNP[17]. Later in 2016, the ABC score was also developed to predict bleeding risk score in patients with AF, and also achieved similar results [18]. Moreover, in 2017, ABC was also developed to predict death in anticoagulated patients with AF, including both clinical information and biomarkers. The model yielded higher AUC than a model based on all clinical variables, both the derivation (0.74 vs. 0.68) and validation cohorts (0.74 vs. 0.67) [19]. Later in 2021, ABC-AF was also validated in patients not receiving oral anticoagulation and yielded similar positive results [2].

In 2020, GARFIELD-AF risk model was developed to predict stroke, major bleeding or mortality in AF patients. The model is described as developed using sophisticated statistical modeling techniques, and can be found on https://af.garfieldregistry.org/garfield-af-risk-calculator. It achieved a higher score in stroke and mortality prediction compared with CHA_2_DS_2_-VASc and a higher score than HAS-BLED score in predicting major bleed events across all risk groups [1]. The model was then externally validated in ORBIT-AF cohort, and the discriminatory value was still superior than CHA_2_DS_2_-VASc score.

In 2022, the Fushumi AF registry, a cohort from Fushumi, Japan, was used to learn predictors of ischemic events in patients with AF. Apart from biological and past medical and treatment history, the model also used medication, blood test, and echocardiogram data. The model was able to have an increased performance compared to CHA_2_DS_2_-VASc with an AUC of 0.72 vs 0.62 [26].

Also in 2022, using the same Fushimi AF registry, another machine learning risk model was developed to predict incident heart failure in patients with AF. The model outperformed Framingham risk score with an AUC of 0.75 vs 0.67 [16]. However, it is to note that the Framingham risk score is not specifically tailored to predict heart failure events in an AF specific population.

In 2024, another model was developed to predict 1-year stroke risk in an Indian population, and achieved an AUC of 0.82 compared with CHA_2_DS_2_-VASc 0.67. However, when externally validated in another Asian cohort, the model only achieved an AUC of 0.67 while CHA_2_DS_2_-VASc achieved 0.62 [6].

More recently, a meta-analysis evaluated 13 studies on stroke prediction in patients with AF using machine learning methods. The mean AUC across studies was 0.73, with models such as XGBoost and logistic regression achieving higher performance, while neural networks showed comparatively lower AUC values [14].

Within the prediction of AF-related outcomes, stroke is the most extensively studied, given the substantial increase in stroke risk associated with AF. Other predictive models have focused on outcomes such as bleeding or all-cause mortality, while less attention has been given to events like acute coronary syndrome, cardiovascular death, and hospitalizations.

## 3 Dataset

The dataset comes from anonymised electronic health records (EHRs) of patients followed at Unidade Local de Saúde de Matosinhos (ULSM) and spans approximately 25 years, including patients over 40 years old who were diagnosed with AF between 1 January 2012 and 31 December 2021. The dataset comprises a population of 7203 patients and 167 features encompassing demographics, clinical features (pertaining to laboratory, pharmaceutical, and surgical acts), and the temporal context of the previous electronic registry.

The study was approved by the Ethical Committee and Data Protection Officer of ULSM. The original data was de-identified according to HIPAA Safe Harbour Method with noise added to all variables.

Table 1 presents key features of the dataset, excluding several binary features associated with the presence of the following comorbidities: chronic obstructive pulmonary disease, myocardial infarction or unstable angina, type 1 diabetes mellitus, type 2 diabetes mellitus, and valvular heart disease. Additionally, there are several outcomes: acute coronary syndrome, arterial embolism, stroke, inpatient visit, heart failure hospitalization, cardiovascular death, and all-cause death.

**Table 1:** Some selected variables of the USLM dataset. Binary features representing comorbidities and dispensed medications are not listed for simplicity’s sake.

| Feature Name | Description | Data Type | Units |
| --- | --- | --- | --- |
| index_date | index date (random) | integer | N/A |
| age_hipaa | age (HIPAA compliant) | categorical | years |
| female | female | binary | N/A |
| wgt | weight | float | kg |
| hgt | height | integer | cm |
| bmi | body mass index | float | kg/m<sup>2</sup> |
| sbp | systolic blood pressure | float | mmHg |
| dbp | diastolic blood pressure | float | mmHg |
| tc | total cholesterol | float | mg/dL |
| ldl | LDL cholesterol | float | mg/dL |
| hdl | HDL cholesterol | float | mg/dL |
| glc | glycemia | float | mg/dL |
| tg | triglycerides | float | mg/dL |
| crt | creatinine | float | mg/dL |
| egfr | estimated glomerular filtration rate | float | mL/min/1.73m<sup>2</sup> |
| tsh | thyroid stimulating hormone | float | mIU/L |
| uacr | urine albumin-to-creatinine ratio | float | mg/g |
| a1c | Hba1c (glycated hemoglobin) | float | mmol/mol |
| cancer | cancer | binary | N/A |
| carotd | carotid disease | binary | N/A |
| cd | coronary disease | binary | N/A |
| dlp | dyslipidemia | binary | N/A |
| esrd | end stage renal disease | binary | N/A |
| flt | flutter | binary | N/A |
| hf | heart failure | binary | N/A |
| hta | hypertension | binary | N/A |
| slap | sleep apnea | binary | N/A |
| stk | stroke | binary | N/A |
| tyrd | thyroid disease | binary | N/A |

The dataset also includes binary variables indicating whether a patient is currently on any of the following medications: antiarrhythmics, anticoagulants, angiotensin-aonverting anzyme anhibitor (ACEi), angiotensin receptor brlockers (ARBs), angiotensin receptor-neprilysin inhibitor (ARNi), antiplatelets, beta blockers, calcium channel blockers, digoxin, dipeptidyl peptidase-4 inhibitor (DPP4i), GLP-1 agonists, insulin, ivabradine, loop diuretics, other diuretics, metformin, mineralocorticoid receptor antagonist (MRA), nitrates, sodium-glucose cotransporter-2 inhibitors (SGLT2i), statins, and sulfonylurea. Moreover, the data contains additional binary variables that represent whether a patient had the following interventions before: cardiac surgery, cardiac device, coronary surgery, and percutaneous coronary intervention. To represent smoking, the dataset has a set of binary features: current smoker, former smoker, never smoked, and no information smoker. Some laboratory tests also include an additional feature representing the patient’s initial measurement for that specific exam. These tests are: HbA1c, creatinine, estimated glomerular filtration rate, glycemia, HDL cholesterol, LDL cholesterol, total cholesterol, triglycerides, thyroid simulating hormone, urine albumin-to-creatinine ratio, and international normalized ratio (INR).

Every feature, except for index date, age (HIPAA), and sex (female), has an associated integer day count indicating when the event occurred relative to the AF diagnosis. For comorbidities, the index date defines the diagnosis data. For laboratory tests, it indicates the date of the measurement. For biometric features, it corresponds to the date the information was extracted, while those for medications refer to the last prescription.

To provide a clearer understanding of the dataset and its key variables, descriptive statistics for selected numerical variables, including mean, standard deviation, and proportion of missing values, are presented in Table 2. As shown, LDL cholesterol, HbA1c, TSH, UACR, and eGFR have a relatively high proportion of missing values (*>*30%), which may impact subsequent analyses. Overall, the mean and standard deviation values indicate moderate variability for most anthropometric and biochemical measures. Triglycerides, glycemia, and UACR display relatively large standard deviations, suggesting substantial variability across participants.

**Table 2:** Descriptive statistics of selected numerical variables: mean and missing rate (after outlier removal).

| Variable | Value (mean ± SD) | Missing Values (%) |
| --- | --- | --- |
| Age* | 77.1 ± 11.2 | 0.0% |
| Weight | 74.4 ± 15.4 | 15.0% |
| Height | 162.1 ± 9.6 | 16.5% |
| BMI | 28.3 ± 5.5 | 17.0% |
| SBP | 136.6 ± 18.9 | 12.2% |
| DBP | 77.4 ± 12.0 | 12.2% |
| Total cholesterol | 176.7 ± 41.9 | 14.3% |
| LDL cholesterol | 108.7 ± 35.6 | 31.2% |
| HDL cholesterol | 45.5 ± 13.6 | 15.0% |
| Triglycerides | 116.9 ± 59.1 | 14.6% |
| Creatinine | 1.1 ± 0.6 | 4.8% |
| Glycemia | 127.2 ± 55.9 | 5.0% |
| Hba1c | 6.3 ± 1.2 | 45.6% |
| TSH | 1.9 ± 1.4 | 33.4% |
| UACR | 33.0 ± 57.1 | 55.7% |
| eGFR | 63.7 ± 21.1 | 35.3% |

\* Age is estimated from categorical age groups using midpoints

Tables 3 and 4 present the distribution of the most relevant variables, including demographic and clinical characteristics. For each variable, the table shows the number of subjects in each category, the corresponding percentage of the total population, and the number and percentage of the cases observed in each outcome within that category. The majority of the population is between 70 and 89 years old (64.8%), with a slightly higher proportion of women than men (53.5% women). BMI and weight are higher than reference values from healthy populations. Hypertension is highly prevalent, affecting 81.8% of the population; however, systolic and diastolic blood pressure values appear to be within normal ranges, likely due to the widespread use of antihypertensive medication (91.5%). Total cholesterol and triglycerides are elevated in only a subset of subjects (24.1% and 17.9%, respectively). Nearly 40% of the population is diabetic (37.4%), and most have never smoked (76.0%). Several comorbidities are common: dyslipidemia (64.7%), heart failure (30.5%), valvular heart disease (26.2%), and cancer (22.7%). Other conditions with notable prevalence include myocardial infarction or unstable angina (17.2%), chronic obstructive pulmonary disease (12.5%), thyroid disease (11.4%), coronary heart disease (10.9%), peripheral artery disease (9.1%), and carotid disease (6.3%). Anticoagulant use was limited prior to AF onset (6.2%) and rose following AF diagnosis. The distribution of outcome incidence across population groups is generally consistent with the underlying group sizes.

**Table 3:** Descriptive statistics of the ULSM cohort.

| Risk Factor | Value | Patients<br>(n = 7154) | Stroke/SE<br>(n = 168) | All-cause Death<br>(n = 1546) | CV Death<br>(n = 1280) | HF Hospitalizations<br>(n = 2284) | Inp. Visit<br>(n = 4176) | ACS<br>(n = 306) |
| --- | --- | --- | --- | --- | --- | --- | --- | --- |
| Age | 40-49 | 130 (1.8%) | 2 (1.2%) | 7 (0.5%) | 3 (0.2%) | 16 (0.7%) | 47 (1.1%) | 3 (1.0%) |
|  | 50-59 | 435 (6.1%) | 7 (4.2%) | 30 (1.9%) | 24 (1.9%) | 93 (4.1%) | 204 (4.9%) | 15 (4.9%) |
|  | 60-69 | 1,178 (16.5%) | 30 (17.9%) | 125 (8.1%) | 107 (8.4%) | 275 (12.0%) | 633 (15.2%) | 57 (18.6%) |
|  | 70-79 | 2,184 (30.5%) | 44 (26.2%) | 380 (24.6%) | 322 (25.2%) | 694 (30.4%) | 1267 (30.3%) | 103 (33.7%) |
|  | 80-89 | 2,453 (34.3%) | 73 (43.5%) | 744 (48.1%) | 620 (48.4%) | 936 (41.0%) | 1584 (37.9%) | 109 (35.6%) |
|  | ≥ 90 | 774 (10.8%) | 12 (7.1%) | 260 (16.8%) | 204 (15.9%) | 270 (11.8%) | 441 (10.6%) | 19 (6.2%) |
| Sex | Male | 3330 (46.5%) | 81 (48.2%) | 710 (45.9%) | 582 (45.5%) | 941 (41.2%) | 1885 (45.1%) | 159 (52.0%) |
|  | Female | 3824 (53.5%) | 87 (51.8%) | 836 (54.1%) | 698 (54.5%) | 1343 (58.8%) | 2291 (54.9%) | 147 (48.0%) |
| BMI<br>(kg/m<sup>2</sup>) | <20 | 255 (3.6%) | 8 (4.8%) | 70 (4.5%) | 55 (4.3%) | 66 (2.9%) | 138 (3.3%) | 12 (3.9%) |
|  | 20-<25 | 1497 (20.9%) | 38 (22.6%) | 353 (22.8%) | 298 (23.3%) | 406 (17.8%) | 861 (20.6%) | 57 (18.6%) |
|  | 25-<30 | 2282 (31.9%) | 50 (29.8%) | 458 (29.6%) | 386 (30.2%) | 704 (30.8%) | 1304 (31.2%) | 105 (34.3%) |
|  | ≥ 30 | 2083 (29.1%) | 41 (24.4%) | 383 (24.8%) | 332 (25.9%) | 773 (33.8%) | 1255 (30.1%) | 93 (30.4%) |
| Height<br>(cm) | <156 | 1647 (23.0%) | 44 (46.2%) | 380 (24.6%) | 322 (25.2%) | 609 (26.7%) | 1013 (24.3%) | 67 (21.9%) |
|  | 156-<164 | 1813 (25.3%) | 47 (28.0%) | 391 (25.3%) | 332 (25.9%) | 596 (26.1%) | 1087 (26.0%) | 66 (21.6%) |
|  | 164-<173 | 1767 (24.7%) | 34 (20.2%) | 350 (22.6%) | 291 (22.7%) | 523 (22.9%) | 1004 (24.0%) | 99 (32.4%) |
|  | ≥ 170 | 922 (12.9%) | 13 (7.7%) | 157 (10.2%) | 138 (10.8%) | 234 (10.2%) | 476 (11.4%) | 36 (11.8%) |
| Weight<br>(kg) | <55 | 561 (7.8%) | 19 (11.3%) | 153 (9.9%) | 122 (9.5%) | 156 (6.8%) | 318 (7.6%) | 22 (7.2%) |
|  | 55-<70 | 1910 (26.7%) | 48 (28.6%) | 439 (28.4%) | 368 (28.7%) | 575 (25.2%) | 1119 (26.8%) | 75 (24.5%) |
|  | 70-<85 | 2347 (32.8%) | 49 (29.2%) | 466 (30.1%) | 394 (30.8%) | 747 (32.7%) | 1372 (32.9%) | 115 (37.6%) |
|  | 85-<100 | 1069 (14.9%) | 20 (11.9%) | 190 (12.3%) | 169 (13.2%) | 371 (16.2%) | 629 (15.1%) | 41 (13.4%) |
|  | ≥ 100 | 359 (5.0%) | 6 (3.6%) | 47 (3.0%) | 41 (3.2%) | 133 (5.8%) | 194 (4.6%) | 15 (4.9%) |
| SBP<br>(mmHg) | <100 | 151 (2.1%) | 1 (0.6%) | 34 (2.2%) | 25 (2.0%) | 46 (2.0%) | 86 (2.1%) | 1 (0.3%) |
|  | 100-<120 | 923 (12.9%) | 13 (7.7%) | 217 (14.0%) | 186 (14.5%) | 320 (14.0%) | 527 (12.6%) | 28 (9.2%) |
|  | 120-<140 | 2659 (37.2%) | 56 (33.3%) | 506 (32.7%) | 416(32.5%) | 760 (33.3%) | 1474 (35.3%) | 108 (35.3%) |
|  | 140-<160 | 2023 (28.3%) | 53 (31.5%) | 414 (26.8%) | 355 (27.7%) | 642 (28.1%) | 1196 (28.6%) | 86 (28.1%) |
|  | ≥ 160 | 662 (9.3%) | 25 (14.9%) | 176 (11.4%) | 149 (11.6%) | 262 (11.5%) | 446 (10.7%) | 47 (15.4%) |
| DBP<br>(mmHg) | <70 | 1604 (22.4%) | 32 (19.0%) | 432 (27.9%) | 359 (28.0%) | 608 (26.6%) | 989 (23.7%) | 78 (25.5%) |
|  | 70-<80 | 2037 (28.5%) | 42 (25.0%) | 409 (26.5%) | 342 (26.7%) | 599 (26.2%) | 1179 (28.2%) | 80 (26.1%) |
|  | 80-<90 | 1891 (26.4%) | 45 (26.8%) | 351 (22.7%) | 300(23.4%) | 542 (23.7%) | 1059 (25.4%) | 69 (22.5%) |
|  | 90-<100 | 693 (9.7%) | 15 (8.9%) | 121 (7.8%) | 100 (7.8%) | 220 (9.6%) | 389 (9.3%) | 33 (10.8%) |
|  | ≥ 100 | 194 (2.7%) | 14 (8.3%) | 35.0 (2.3%) | 31 (2.4%) | 62 (2.7%) | 114 (2.7%) | 11 (3.6%) |
| Total<br>cholesterol<br>(mg/dL) | <200 | 4559 (63.7%) | 103 (61.3%) | 1030 (66.6%) | 852 (66.6%) | 1524 (66.7%) | 2694 (64.5%) | 196 (64.1%) |
|  | 200-<240 | 1290 (18.0%) | 35 (20.8%) | 255 (16.5%) | 208 (16.2%) | 415 (18.2%) | 783 (18.8%) | 60 (19.6%) |
|  | ≥ 240 | 434 (6.1%) | 8 (4.8%) | 80 (5.2%) | 70 (5.5%) | 116 (5.1%) | 252 (6.0%) | 23 (7.5%) |
| Triglycerides<br>(mg/dL) | <150 | 4985 (69.7%) | 118 (70.2%) | 1100 (71.2%) | 913 (71.3%) | 1624 (71.1%) | 2941 (70.4%) | 211 (69.0%) |
|  | 150-<200 | 767 (10.7%) | 13 (7.7%) | 148 (9.6%) | 125 (9.8%) | 250 (10.9%) | 455 (10.9%) | 38 (12.4%) |
|  | ≥ 200 | 516 (7.2%) | 13 (7.7%) | 109 (7.1%) | 89 (7.0%) | 175 (7.7%) | 322 (7.7%) | 30 (9.8%) |
| Diabetes<br>mellitus (mg/dL) | Type 1 | 323 (4.5%) | 11 (6.5%) | 89 (5.8%) | 72 (5.6%) | 149 (6.5%) | 221 (5.3%) | 28 (9.2%) |
|  | Type 2 | 2361 (32.9%) | 60 (35.7%) | 564 (36.5%) | 466 (36.4%) | 882 (38.6%) | 1468 (35.2%) | 141 (46.1%) |

**Table 4:**
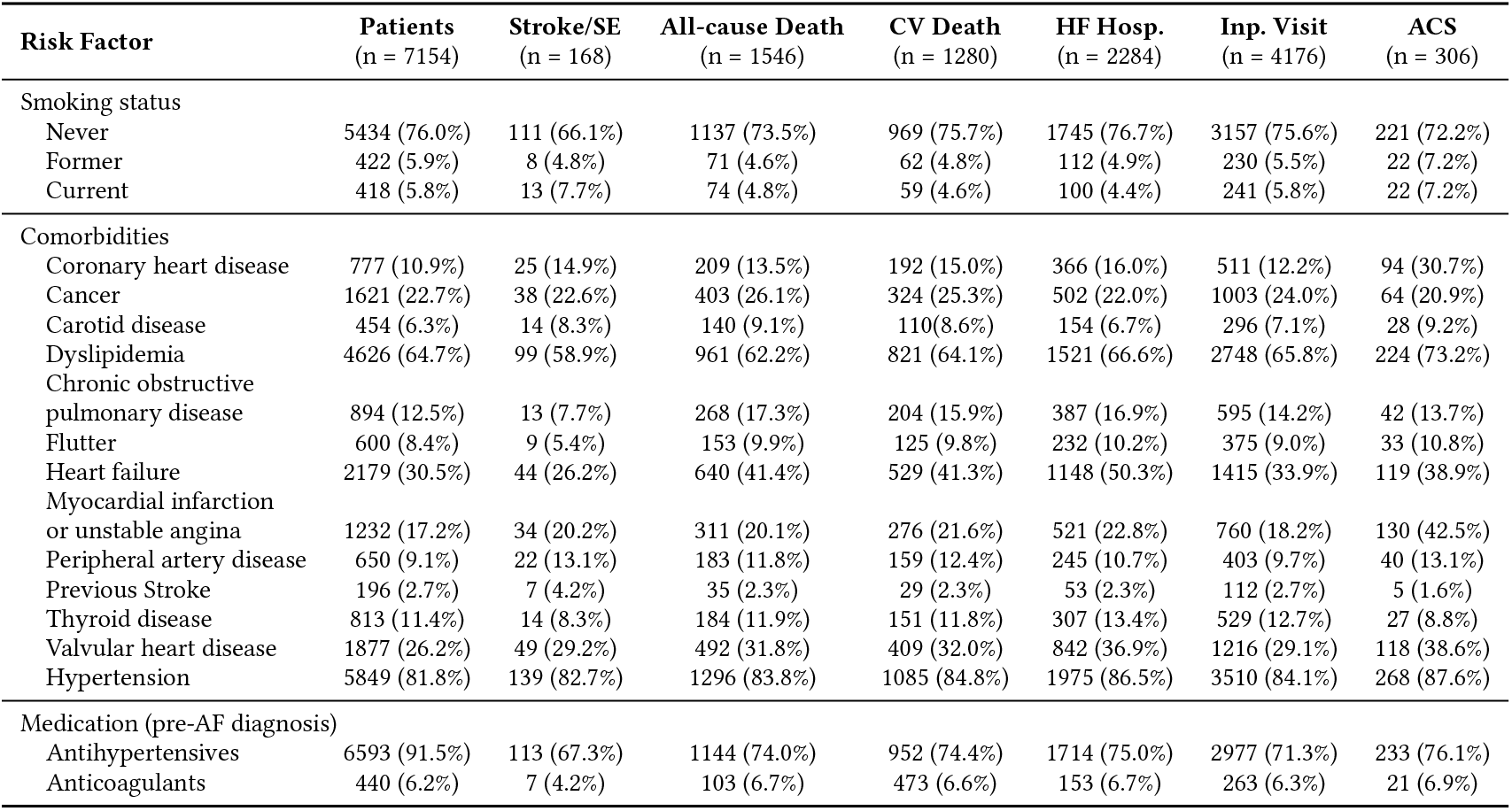
Descriptive statistics of the ULSM cohort.

In Figure 1, the distribution of elapsed time from AF diagnosis to each outcome is shown. Most events occur shortly after diagnosis, with their frequency decreasing progressively over time. Stroke and arterial embolism are shown together due to low occurrence.

**Figure 1:**
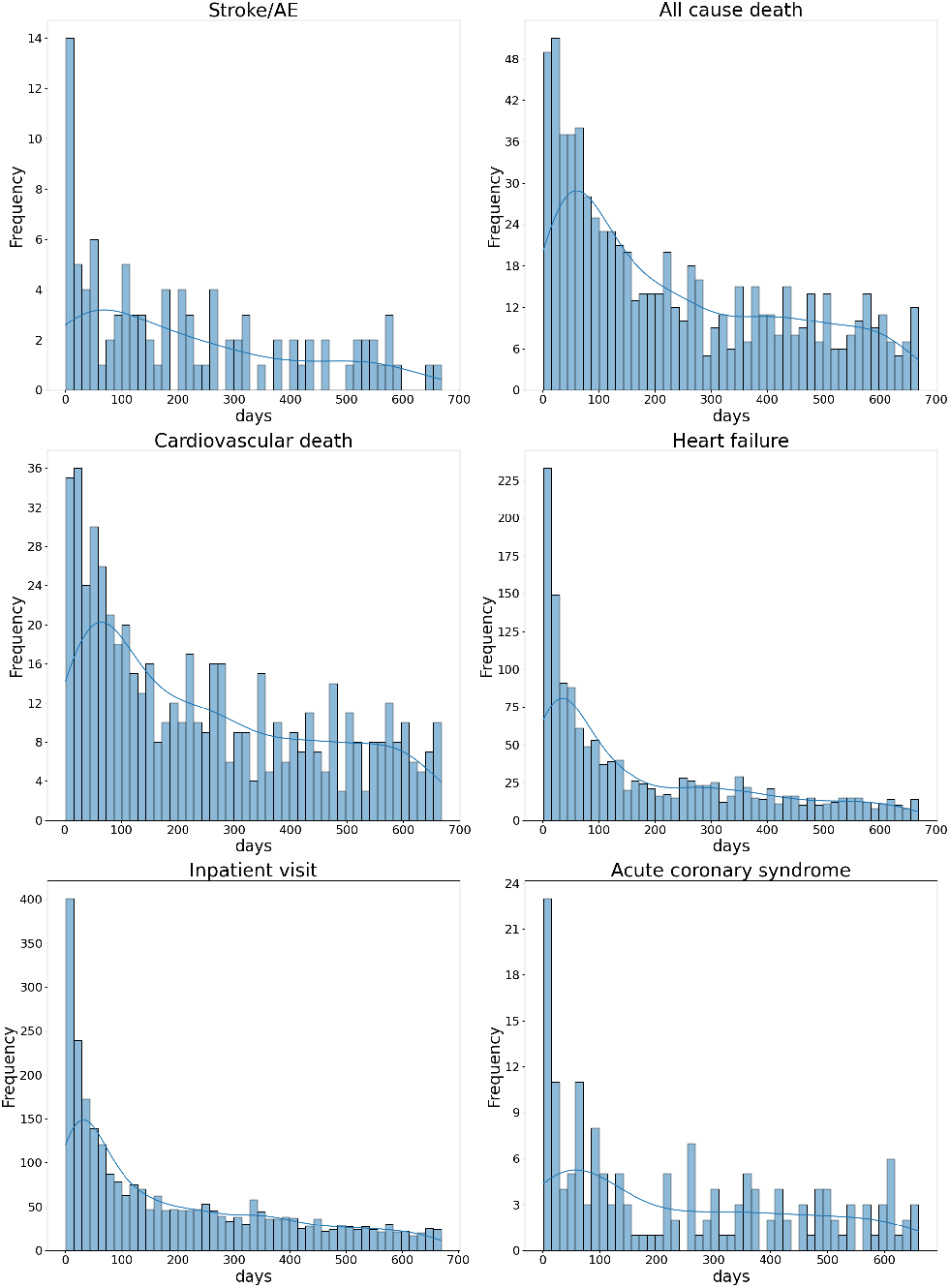
Distribution of elapsed time (in days) from AF diagnosis to outcome per clinical endpoint. Histograms with 14-day bins and truncated at 2 years of follow-up.

## 4 Methods

### 4.1 Data Preprocessing

The study followed a structured data preprocessing and modeling workflow. Data cleaning involved recalculating all time-related variables relative to each patient’s atrial fibrillation diagnosis and excluding individuals under 40 years of age. Outliers were treated through capping and log transformation, and implausible values were corrected. Missing data was handled through targeted removal of incomplete records and mean imputation, after evaluating each variable’s relevance. Constant and redundant features were removed, and new clinically meaningful binary features were engineered, including aggregated indicators capturing the presence of vascular disease, diabetes, kidney function, and medication use. Temporal variables were used to derive longitudinal and delta-based representations to capture within-patient variation over time. Three dataset versions were produced: static, slope-based, and longitudinal. Class imbalance was addressed through resampling methods such as random sampling and SMOTE, depending on the outcome and model. Finally, standardization using the StandardScaler was applied to scale-sensitive algorithms (e.g., logistic regression, Naïve Bayes, multilayer perceptron), ensuring that the training and test sets remained properly separated.

### 4.2 Classical Risk Calculators

To evaluate the performance of classical risk calculators within the AF cohort, two representative models were selected: CHA_2_DS_2_ -VASc, which follows a point-based system, and GARFIELD-AF, which is based on Cox regression. Both models were selected for their ability to predict stroke, with GARFIELD-AF also capable of predicting mortality in AF cohorts. These scores serve as a baseline for comparison with subsequently developed machine learning models, as they are specifically tailored for these comorbidities in AF cohorts.

To access stroke risk in the cohort CHA_2_DS_2_-VASc score is further calculated, estimating stroke/SE risk over a 1-year period. CHA_2_DS_2_-VASc is a risk-stratification score ranging from 0 to 9, depending on the number and weight of the score’s risk components. Table 5 shows each risk factor and the corresponding score.

**Table 5:** CHA_2_DS_2_-VASc risk factors and corresponding score.

| Risk Factor | Score (cf. [25]) |
| --- | --- |
| Congestive heart failure (C) | 1 |
| Hypertension (H) | 1 |
| Age 75+ years (A) | 2 |
| Diabetes mellitus (D) | 1 |
| Stroke (S) | 2 |
| Vascular disease (V) | 1 |
| Age 65-74 years (A) | 1 |
| Sex category - female (Sc) | 1 |

\* If a woman is assigned only one point due to sex category, her score should be adjusted to zero.

The ULSM dataset contains all necessary information to compute CHA_2_DS_2_ -VASc score. Data on congestive heart failure (equivalent to heart failure), hypertension, diabetes mellitus, stroke, and sex are directly available. Vascular disease was derived in the data preprocessing. Age was calculated using the middle value of the category interval: patients aged 60–69 were assigned a score of 1 (using 65 as the representative age), while those aged 70–79 or older were assigned a score of 2.

The GARFIELD-AF score was calculated for all patients in the cohort to evaluate their risk of stroke and all-cause mortality. Calculation of the GARFIELD-AF score requires the following variables: age, sex, ethnicity, diastolic blood pressure, pulse, history of heart failure, vascular disease, prior stroke, history of bleeding, diabetes, moderate-to-severe chronic kidney disease, dementia, current smoking, and use of vitamin K antagonist (VKA) or non–vitamin K antagonist oral anticoagulants (NOACs). Among these, sex, diastolic blood pressure, history of heart failure, prior stroke, current smoking, and chronic kidney disease were directly available from the dataset. Vascular disease and diabetes were derived during data preprocessing. Age was estimated as the midpoint of each age category, similarly to CHA_2_DS_2_ -VASc score. Variables not captured in the dataset were handled with assumptions: pulse was set at 80 bpm, and patients were assumed to have no history of bleeding or dementia. Regarding anticoagulant use, VKA treatment was assumed absent, and NOAC treatment was considered present for patients recorded as taking any anticoagulant. The score for each outcome was then computed using the original regression coefficients, summarized in Table 6, together with the Cox proportional hazards formulas for all-cause mortality, ischemic stroke and systemic embolism^1^

**Table 6:** GARFIELD-AF coefficients for 6-month all-cause mortality and 1-year ischemic stroke/systemic embolism.

| Variable | All-cause death $\beta$ | Stroke/SE $\beta$ |
| --- | --- | --- |
| Female sex | -0.306202287 | - |
| Heart failure | 0.693789082 | 0.233182644 |
| Vascular disease | 0.306120964 | 0.197919709 |
| Prior stroke | 0.265852980 | 0.800863063 |
| History of bleeding | 0.385407386 | 0.298839670 |
| Diabetes | 0.280133213 | 0.211995445 |
| Moderate-to-severe CKD | 0.377903886 | 0.349516938 |
| Dementia | 0.489453313 | 0.513221391 |
| Current smoking | 0.345481149 | 0.478831506 |
| OAC treatment: NOAC | -0.414591263 | -0.572199357 |
| OAC treatment: VKA | -0.185935610 | -0.352373263 |
| Ethnicity: Hispanic/Latino | 0.157023564 | - |
| Ethnicity: Asian | -0.609609055 | - |
| Ethnicity: Black/Other | 0.375675102 | - |
| (Age-65) (if $\leq 65$) | 0.031050027 | 0.039138147 |
| (Age-65) (if $> 65$) | 0.064594824 | - |
| (Weight-75) (if $\leq 75$) | -0.021535182 | - |
| (Pulse-120) (if $\leq 120$) | 0.007678035 | - |
| (Diastolic BP-80) (if $\leq 80$) | -0.019304333 | - |
| (Diastolic BP-80) (if $> 80$) | - | 0.015900160 |

### 4.3 Machine Learning Predictors

To predict the outcomes, a set of well-established machine learning models was implemented. These included Naïve Bayes [9], Logistic Regression [7], Decision Tree [4], Random Forest [3], XGBoost [5], and a Multi-Layer Perceptron (MLP) [28]. This selection encompasses classical statistical methods, modern ensemble and neural network approaches, allowing for a comprehensive evaluation of predictive performance across different modeling strategies.

Each model was trained for every outcome variable at the 6-month time horizon, except for stroke combined with arterial embolism, which was trained using data from the full 10-year cohort due to extreme class imbalance. Models were trained separately for each dataset type, static, slope-based, and longitudinal, and both independently for each outcome, as well as jointly in a multi-target framework.

Robustness was ensured through 5-fold cross-validation, with model hyperparameters optimized via Bayesian optimization using the F_2_ score as the objective, and final selection was also guided by the F_2_ score during sampling. Using the F-measure with *β* = 2 gives greater weight to recall, ensuring higher sensitivity to false negatives compared to false positives. For each outcome, the best-performing predictor was identified through rank fusion of F_1_, F_2_, and AUC. The best-performing model was subsequently used to assess performance across all time horizons and subjected to explainability analysis using SHAP-based feature importance.

## 5 Results and Discussion

### 5.1 All-Cause Death

Figure 2 shows the distribution of GARFIELD-AF predictions at 6 months in the cohort, indicating the presence of several patients at high risk (≥ 10%) of mortality within the next six months. An AUC of 0.72 demonstrates good discriminatory ability, indicating that the model can reliably differentiate between patients who did and did not experience all-cause mortality within six months.

**Figure 2:**
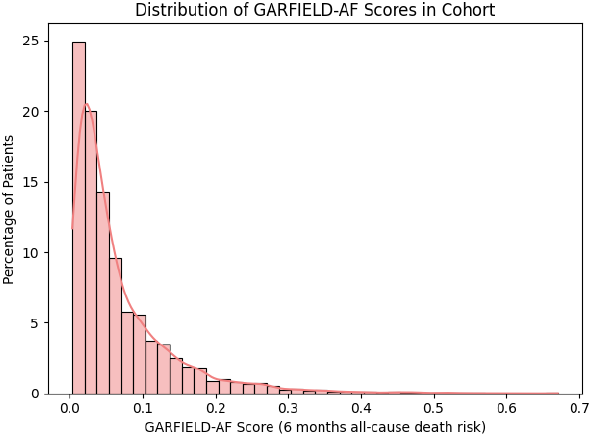
Distribution of GARFIELD-AF scores in the cohort for predicting death at 6 months.

Table 7 presents the performance of the ML models in predicting all-cause death at 6 months. XGBoost, Random Forest, and Logistic Regression achieved the best overall performance, combining high sensitivity with strong precision. The remaining models showed inferior performance: Naive Bayes showed higher sensitivity but at the expense of precision, while the MLP and Decision Tree exhibited generally poor performance. Incorporating slope features did not improve predictive ability, whereas including the full set of longitudinal features enhanced model performance, indicating that these longitudinal features contain valuable predictive information.

**Table 7:**
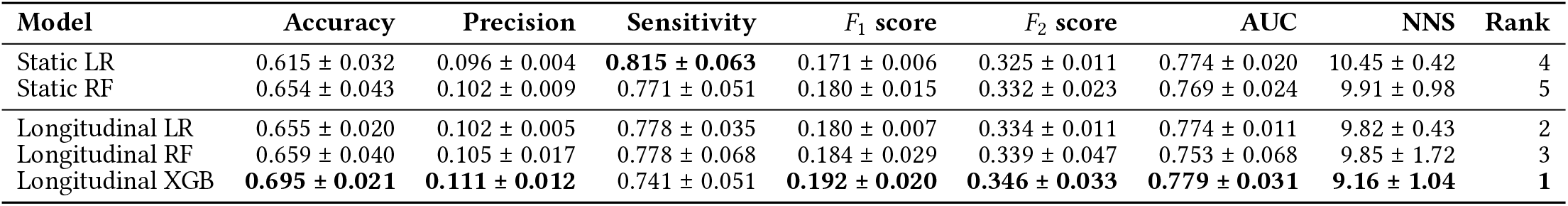
Performance of top performance machine learning models in predicting all-cause death, reported as mean ± standard deviation for accuracy, precision, sensitivity, F_1_ score, F_2_ score, AUC, and NNS. The best model is represented in bold.

The best-performing model was the longitudinal XGBoost, and its AUC for 6-month all-cause death risk (0.779) exceeded that of GARFIELD-AF (0.715), showing the efficacy of machine learning predictors. Compared with the best-performing static model (static LR), the longitudinal XGBoost model incorporating time-aware features demonstrated statistically significant improvements in both F_2_ score (*p* = 4.17*e* − 5) and AUC (*p* = 1.28*e* − 7).

The longitudinal XGBoost has its feature importance presented in Figure 3. Age emerges as the strongest predictor, consistent with its well-established role as a major risk factor. A clinical history of cancer, heart failure, and COPD also contributed to increased risk. BMI was likewise identified as a relevant predictor within the model. In addition, low HDL cholesterol and elevated HbA1c levels (reflecting prolonged hyperglycemia) were linked to increased risk.

**Figure 3:**
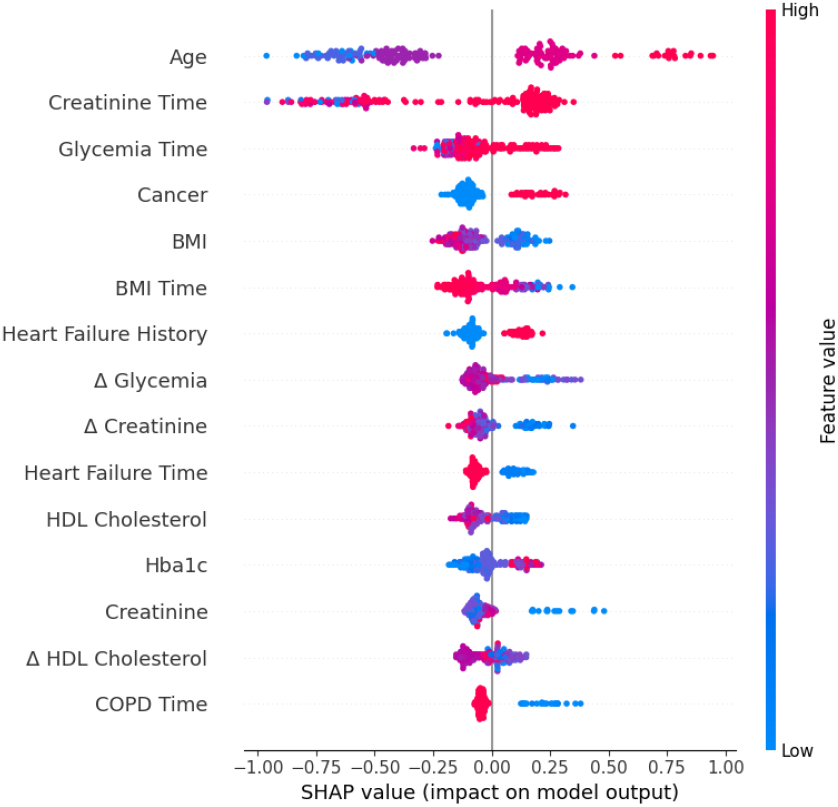
SHAP value of longitudinal XGBoost model on predicting all-cause death at 6 months.

Among the slope-based features, temporal variation in creatinine, glycemia, and HDL cholesterol appeared particularly relevant, suggesting that changes over time in these biomarkers carry predictive information beyond single measurements. The longitudinal (temporal) features overall showed high importance within the model. Time-related variables associated with binary indicators enabled the model to capture both the occurrence and timing of clinical events; notably, the timing and presence of measurements for creatinine, digoxin use, BMI, glycemia, and loop diuretics were influential. However, interpretation of these temporal features remains complex.

To identify the most influential predictors in a static model, static Logistic Regression was selected, and its feature importance is illustrated in Figure 4. The results show features more aligned with medical knowledge. Established features, including weight, HDL cholesterol, systolic blood pressure (SBP), estimated glomerular filtration rate (eGFR), and total cholesterol, consistently emerged as important predictors. Weight reflected the obesity paradox, with higher values associated with lower risk. Low HDL cholesterol increased risk, as expected, while reduced eGFR indicated kidney dysfunction. Interestingly, lower total cholesterol was associated with a higher risk, contrary to expectations but potentially explained by statin use. Elevated SBP was linked to greater risk of death, consistent with established medical knowledge, whereas low diastolic blood pressure appeared to increase risk, a surprising finding, as it is not typically considered a strong risk factor in these populations. Age also contributed modestly, with higher values associated with increased risk, as expected.

**Figure 4:**
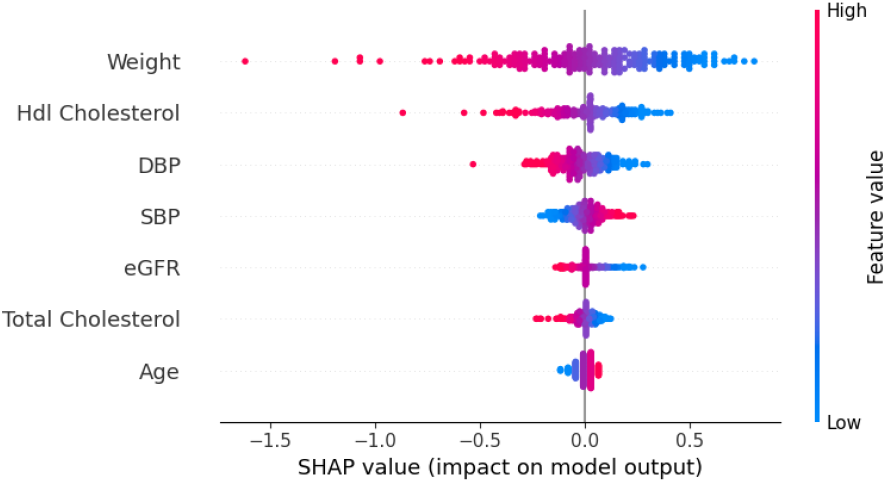
SHAP value of static Logistic Regression model on predicting all-cause death at 6 months.

The longitudinal XGBoost model was then further evaluated across different time intervals (Table 8). The model performance generally improves as the time horizon increases, reflecting the larger number of positive cases; however, the highest AUC is observed at 3 months.

**Table 8:** Performance of longitudinal XGBoost model in predicting all-cause death at different time ranges, reported as mean ± standard deviation for F_1_ score, F_2_ score, and AUC. The best model is represented in bold.

| Interval | $F_1$ score | $F_2$ score | AUC |
| --- | --- | --- | --- |
| 1 month | 0.061 ± 0.019 | 0.095 ± 0.031 | 0.722 ± 0.049 |
| 3 months | 0.165 ± 0.025 | 0.279 ± 0.047 | **0.803 ± 0.030** |
| 6 months | 0.192 ± 0.020 | 0.346 ± 0.033 | 0.779 ± 0.031 |
| 1 year | 0.243 ± 0.022 | 0.412 ± 0.033 | 0.768 ± 0.038 |
| 2 years | **0.312 ± 0.014** | **0.478 ± 0.022** | 0.753 ± 0.013 |

### 5.2 Stroke and Systemic Embolism

The CHA_2_DS_2_-VASc score was developed to estimate 1-year stroke or systemic embolism risk in patients with atrial fibrillation, with traditional risk categories defined as 0 for low risk, 1 for intermediate risk, and ≥ 2 for high risk [25]. According to the 2020 ESC guidelines for the diagnosis and management of AF, risk stratification should be interpreted in a sex-specific manner: low risk (score = 0 in men, or 1 in women), for whom antithrombotic therapy is generally not recommended; intermediate risk (score = 1 in men, or 2 in women), for whom oral anticoagulation (OAC) may be considered; and high risk (score ≥ 2 in men, or ≥ 3 in women), for whom OAC is recommended [21].

In our cohort, the distribution of CHA_2_DS_2_-VASc scores is shown in Figure 5, with a mean of 4.09 and a standard deviation of 1.44. The score achieved an area under the curve (AUC) of 0.588. Most patients had elevated scores, with approximately 94% classified as high risk, 5% as intermediate risk, and 1% as low risk according to the ESC 2020 thresholds. These results are consistent with the presence of a high-risk, frail population and indicate that our dataset contains strong predictors of stroke and systemic embolism. The ROC curve results were also favorable, with the CHA_2_DS_2_- -VASc score achieving an AUC comparable to that of its original derivation cohort (0.588 vs. 0.606).

**Figure 5:**
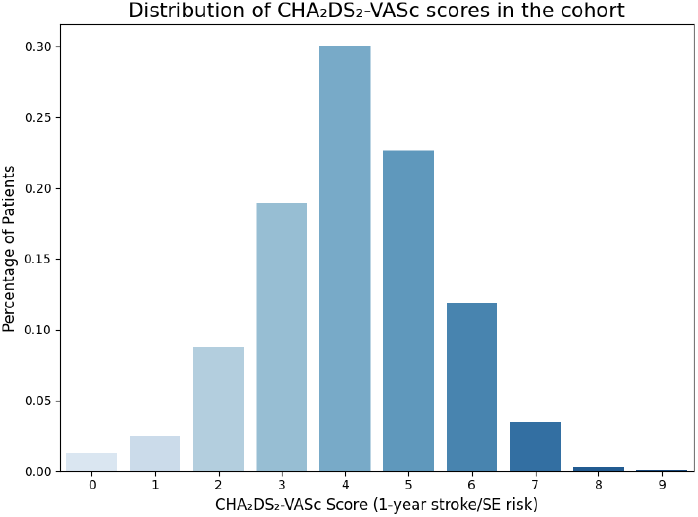
Distribution of CHA_2_DS_2_-VASc scores in the cohort for predicting stroke or systemic embolism at 1 year.

Figure 6 shows the distribution of GARFIELD-AF risk estimates for 1-year ischemic stroke or systemic embolism across the cohort. Although GARFIELD-AF has not defined formal risk categories, higher deciles of predicted risk correspond to increased observed stroke incidence [1]. GARFIELD-AF also demonstrates superior predictive performance compared with CHA_2_DS_2_-VASc, achieving a higher AUC both in the derivation cohort (0.65 vs. 0.59) and in our AF cohort (0.633 vs. 0.588). In this task, an AUC of 0.63 in a validation cohort represents a strong performance, demonstrating the efficacy of GARFIELD-AF in predicting stroke and systemic embolism. It is also noteworthy that several variables required to calculate GARFIELD-AF, such as pulse, ethnicity, bleeding history, dementia, and type of anticoagulant (NOAC vs. VKA), had to be approximated or inferred from our available dataset, which may have affected the predictive accuracy of the score. Nevertheless, the good AUCs observed for both CHA_2_DS_2_-VASc and GARFIELD-AF demonstrate the strength of these predictors, and indicate that the dataset provides reliable indicators of stroke and arterial embolism risk.

**Figure 6:**
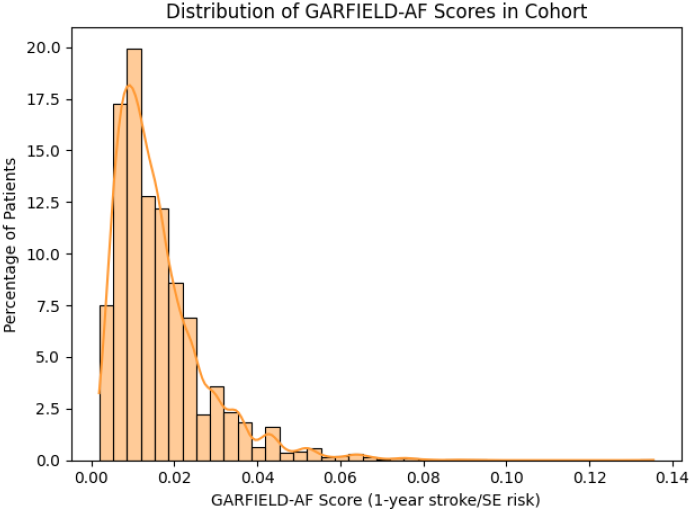
Distribution of GARFIELD-AF scores in the cohort for predicting stroke or systemic embolism at 1 year.

Table 9 summarizes the performance of the machine learning models in predicting stroke or arterial embolism over a 10-year period. The best-performing model was Logistic Regression (LR), with the Multi-Layer Perceptron (MLP) and XGBoost trailing behind. Among the models, the static XGBoost achieved the highest precision, while the slope-based LR attained the highest *F*_1_ and *F*_2_ scores, reflecting a strong balance between precision and sensitivity. Additionally, LR achieved the highest AUC (0.634) and the lowest NNS (29.24), highlighting its superior overall performance.

**Table 9:** Top performance machine learning models in predicting stroke/SE, reported as mean ± standard deviation for accuracy, precision, sensitivity, F_1_ score, F_2_ score, AUC, and NNS. The best models in each metric are highlighted in bold.

| Model | Accuracy | Precision | Sensitivity | $F_1$ score | $F_2$ score | AUC | NNS | Rank |
| --- | --- | --- | --- | --- | --- | --- | --- | --- |
| Static LR | 0.522 ± 0.017 | 0.030 ± 0.001 | **0.640 ± 0.028** | 0.056 ± 0.003 | 0.125 ± 0.006 | 0.605 ± 0.022 | 33.93 ± 1.76 | 3 |
| Static MLP | **0.859 ± 0.018** | **0.036 ± 0.012** | 0.205 ± 0.076 | 0.061 ± 0.021 | 0.106 ± 0.037 | 0.574 ± 0.043 | 32.59 ± 15.23 | 4 |
| Slope-based LR | 0.612 ± 0.031 | 0.034 ± 0.002 | 0.603 ± 0.059 | **0.065 ± 0.004** | **0.140 ± 0.008** | **0.634 ± 0.022** | **29.24 ± 1.70** | **1** |
| Slope-based MLP | 0.856 ± 0.014 | 0.035 ± 0.008 | 0.205 ± 0.047 | 0.060 ± 0.013 | 0.104 ± 0.023 | 0.575 ± 0.033 | 29.91 ± 6.45 | 5 |
| Longitudinal LR | 0.774 ± 0.008 | 0.034 ± 0.008 | 0.338 ± 0.071 | 0.063 ± 0.014 | 0.122 ± 0.027 | 0.611 ± 0.032 | 30.65 ± 7.45 | 2 |

The Naive Bayes demonstrated high sensitivity, while the remaining models showed comparatively limited performance, generally characterized by low precision and sensitivity. Incorporating slope and other longitudinal features enhanced model performance, highlighting the added value of temporal variables in predicting stroke or systemic embolism.

The analysis indicates that the features with the highest relevance in the model and associated with an increased risk of stroke or systemic embolism include body mass index, diastolic blood pressure, eGFR, HDL cholesterol, height, LDL cholesterol, and weight. Most of these variables are well established clinical risk factors. However, height is not traditionally recognized as a risk factor for stroke or systemic embolism. Nevertheless, previous studies have reported a putative association between stature and cardiovascular disease or stroke [27].

The model was subsequently trained across additional time horizons, including 1 year, to assess its performance and compare it with CHA_2_DS_2_-VASc and GARFIELD-AF. The results for these time ranges are presented in Table 10. Model performance increases from 6 months to 1 year and is similar at 1 year and 2 years; however, the highest AUC is observed at 1 year. The 1-month and 3-month horizons were excluded, as the degree of class imbalance prevented the generation of reliable predictions.

**Table 10:** Performance of slope-based Logistic Regression model in predicting stroke and artery embolism at different time ranges, reported as mean ± standard deviation for F_1_ score, F_2_ score, and AUC.

| Interval | $F_1$ score | $F_2$ score | AUC |
| --- | --- | --- | --- |
| 6 months | $0.020 \pm 0.007$ | $0.046 \pm 0.017$ | $0.582 \pm 0.078$ |
| 1 year | $0.034 \pm 0.014$ | $0.076 \pm 0.031$ | **$0.651 \pm 0.098$** |
| 2 years | $0.034 \pm 0.004$ | $0.076 \pm 0.010$ | $0.562 \pm 0.055$ |

The static Logistic Regression model achieved only modest predictive performance, with very low precision and sensitivity. Nevertheless, its AUC for 1-year stroke risk (0.651) exceeded that of CHA_2_DS_2_-VASc (0.588) and GARFIELD-AF (0.633), outperforming the classical risk calculators.

## 6 Clinical Decision Support Tool

To integrate the final models, a clinical decision support tool was developed. It comprises two core components: a backend API and a graphical user interface (GUI). Together, they enable seamless interaction between predictive models and end users.

The API enables integration with existing clinical systems and workflows, offering standardized access to the predictive models and front-end interface. It was implemented using FastAPI, a modern Python web framework chosen for performance and scalability.

Key endpoints such as /predict accept patient data in JSON format, process it through the trained machine learning models, and return predictions with associated probabilities. This design decouples the computational backend from the user interface, allowing multiple clients (e.g., the Dash app, hospital systems, or third-party services) to securely access the prediction service.

Responses are returned in JSON, and the system runs on an ASGI server (uvicorn) that can be deployed standalone or containerized. This modular architecture enhances maintainability, enabling model updates without modifying the interface or external integrations.

The GUI serves as the front end of the tool, allowing clinicians to input patient data and visualize predictions. Built with Dash, it provides interactive forms for variables such as age, weight, blood pressure, and laboratory values. Future versions will support dynamic form customization to handle missing or optional inputs.

Predictions are displayed through clear visualizations that position the patient’s data within the training cohort, improving interpretability. When data is imputed, the GUI communicates with the backend API, retrieves predictions, displaying results in real time. By separating the interface from the API, the system maintains flexibility, visual elements and layouts can evolve independently of the underlying models. Although the current version supports predictions for cardiovascular death, the framework is designed to extend to additional outcomes, including AF onset and related complications.

Overall, the prototype demonstrates that predictive modeling can be effectively integrated into a practical, user-friendly clinical tool. Its modular, extensible design ensures that future iterations can expand both functionality and usability while remaining compatible with clinical workflows.

Figure 7 presents the graphical user interface of the tool, showing how patient data is entered and processed through the API. The source code is available at GitHUB via https://github.com/rique-git/AF-DETECT.

**Figure 7:**
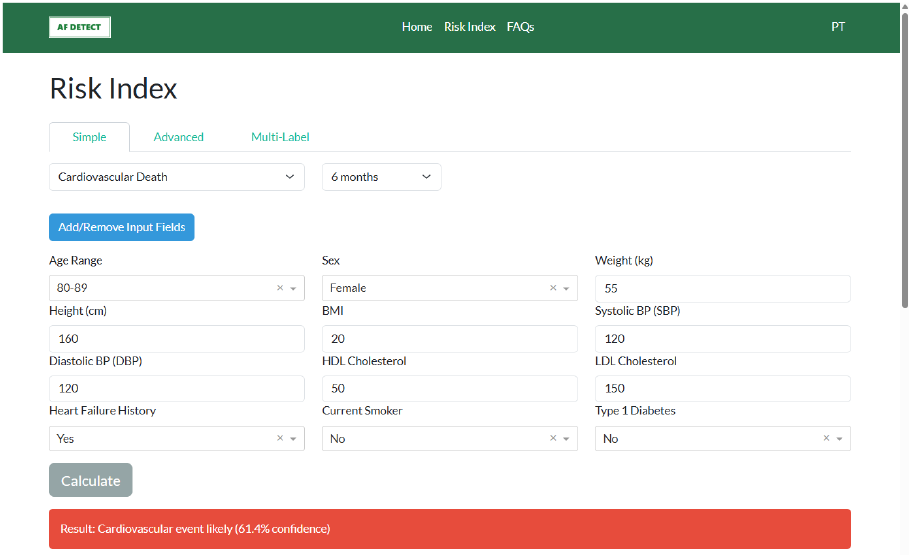
Screenshot of the prototype interface of the medical prediction tool for predicting cardiovascular death at 6 months using key clinical variables.

## 7 Conclusion

### 7.1 Concluding Remarks

This work addressed the challenge of prognosticating atrial fibrillation related outcomes by: (1) developing predictive models for clinically relevant endpoints, and (2) designing a prototype clinical decision-support tool. Together, these efforts aim to improve AF management in primary care.

The predictive models achieved solid performance, often surpassing classical risk calculators for stroke/SE, and all-cause mortality, and performed robustly across different outcomes and time horizons. Integrating diverse diagnostic, clinical, and behavioral data, along with temporal information, enabled a more holistic view of patient risk. Including timing-related features, such as diagnosis and medication events, consistently improved model performance by capturing progression patterns.

Across outcomes, several recurring associations emerged. Height showed a consistent inverse relationship with risk, possibly reflecting early-life social or physical factors influencing cardiovascular health. The “obesity paradox” was also observed: patients with lower BMI tended to have higher risk, likely due to frailty or confounding by illness. Lipid patterns showed that lower LDL and total cholesterol were often linked to higher risk, likely reflecting wide-spread statin use in the cohort. Blood pressure variables provided further insight, high systolic pressure increased risk as expected, but lower diastolic pressure was also associated with poorer outcomes, potentially due to medication effects. Notably, hypertension itself rarely appeared as important, likely because it was nearly universal in the population and thus lost predictive value.

Certain comorbidities, such as diabetes, did not consistently appear as predictors, while medication use (e.g., insulin, digoxin, loop diuretics) often did, highlighting how treatment practices shape model interpretation. Clinical variables like eGFR and age contributed meaningfully to predictions, though sex was significant only for heart failure hospitalization. These findings emphasize that in a heavily medicated AF cohort, traditional risk factors often behave differently, providing valuable insight into the complex interplay between treatment and risk.

While temporal models dominated, interpreting temporal features remains challenging. Some variables, such as “nitrates time,” likely represent medication use rather than true temporal dynamics. Others, linked to clinical activity, may reflect healthcare utilization patterns rather than underlying pathology, an important consideration when interpreting predictive importance.

Overall, this work highlights the potential of time-aware ML for AF risk prediction, uncovers meaningful population-specific patterns, and lays groundwork for clinical implementation. The results show initial promise towards risk prediction, aiding the prognostication of patients with atrial fibrillation.

### 7.2 Limitations and Future Work

Some limitations should be acknowledged. The dataset is collected from a cohort study monitored at ULS Matoshinhos, which can hamper the generalizability of the predictors towards other demographies and populations. Important information such as AF type, pulse, dementia status, anticoagulant class (NOAC vs. VKA), cardiac murmurs, and ethnicity are not included. Additionally, data on bleeding history, liver health, and alcohol use, which is key for predicting bleeding risk, are unavailable.

Future work should include richer electronic health record integration and expand to include control populations to enable AF incidence prediction. Access to electrocardiograms (ECGs), whether as raw signals or extracted features, would provide valuable complementary data, enhancing model robustness and interpretability.

Methodologically, models were designed for longitudinal data but not fully optimized for its structure. Future studies could explore deep learning architectures that capture variable–timestamp relationships and irregular sampling patterns. Improved feature engineering, such as clinically informed thresholds for lab values like eGFR or creatinine, could yield more interpretable and accurate predictions. Multi-output learning also warrants further exploration, especially for related outcomes such as cardiovascular and all-cause death. Threshold tuning to optimize specific clinical metrics (*F*_1_, *F*_2_, or cost-sensitive loss functions) could further enhance applicability.

Future work should further assess predictors on independent cohorts using external validation before consider real-world deployment. Finally, the prototype interface should be refined through iterative design with clinicians to improve usability and integration into clinical workflows. With continued development, this system could serve as a practical tool supporting evidence-based, data-driven AF management in primary care settings.

## Data Availability

Access to all data in the present study should be requested to Unidade Local de Saude de Matosinhos (ULSM), together with a description of the target aims and any additional documentation deemed relevant for ethics approval.

## Acknowledgments

The authors thank Unidade Local de Saúde de Matosinhos (ULSM) for providing the anonymized data. Henrique Anjos, Ana Lebreiro, Cristina Gavina, Rui Henriques, and Rafael S. Costa

## Ethics declarations

Ethical approval was obtained from the Unidade Local de Saúde de Matosinhos (ULSM) committee for the analysis of the anonymized dataset. The authors declare that they have no competing interests.

## Footnotes

1 GARFIELD-AF all-cause mortality = 1 − 0.987921904^exp(∑*βX*)^ GARFIELD-AF ischemic stroke/SE = 1 − 0.9925445321^exp(∑*βX*)^ where *β* is the regression coefficient associated with each risk factor, *X* is the value or presence of each corresponding risk factor, with the respective adjustments. All-cause mortality equation is for estimating risk at 6 months, and the ischemic stroke/SE formula is for estimating risk at 1 year. The original GARFIELD-AF equations are expressed as percentages and are therefore multiplied by 100. Other GARFIELD-AF formulas can be found at https://af.garfieldregistry.org/garfield-af-risk-calculator.

